# Therapeutic effects of *Rosa Canina, Urtica Dioica* and *Tanacetum Vulgare* Herbal Combination in Treatment of Tinnitus Symptoms; A Double-blind Randomized Clinical Trial

**DOI:** 10.1101/2022.03.04.22271916

**Authors:** Mohammad Hossein Khosravi, Amirhomayoun Atefi, Afsaneh Mehri, Fatemeh Sodeifian, Jaleh Yousefi, Ali Bagheri Hagh, Saeed Sohrabpour, Fatemeh Kazemi, Mohammad Ajalloueian, Masoumeh Saeedi

**Affiliations:** Department of Research, Arka Education and Clinical Research Consultants, Tehran, Iran; Student Research Committee, School of Medicine, Guilan University of Medical Sciences, Rasht, Iran; Faculty of Pharmacy, Tehran Medical Sciences, Islamic Azad University, Tehran, Iran; Student research Committee, School of Medicine, Shahid Beheshti University of medical science, Tehran, Iran; Department of Otorhinolaryngology, Faculty of Medicine, Baqiyatallah University of Medical Sciences, Tehran, Iran; Otorhinolaryngology Research Center, Tehran University of Medical Sciences, Tehran, Iran; Student Research Committee, Qazvin University of Medical Sciences, Qazvin, Iran; New Hearing Technologies Research Center, Baqiyatallah University of Medical Sciences, Tehran, IR Iran

**Keywords:** Rosa Canina, Urtica Dioica, Tanacetum Vulgare, Tinnitus and Neurotec

## Abstract

**Background:** Tinnitus is defined as the perception of sound in the ear or head in the absence of an external stimulus for which we have no definite treatment. Neurotec® is a medication of herbal origin with IFDA approval. Previous studies showed the neuroprotective effect of Neurotec®. In this study we evaluated the effectiveness of Neurotec in improving tinnitus symptoms.

**Methods:** This double-blind randomized clinical trial was performed on patients with tinnitus. Patients received Neurotec 100 mg capsules (BID) or placebo for three months. Pure tone audiometry (PTA) was measured at 0.5, 1, 2, 4 and 6 KHz frequencies. Using a Tinnitus Handicap Inventory (THI) questionnaire, tinnitus loudness, daily annoyance, daily life or sleep disturbance, daily perception and mood alteration were evaluated.

**Results:** Finally, 103 (69 male and 34 female) patients with a mean age of 51.33±13.91 years were analyzed. There was no significant difference between the intervention (n=53) and the control group (n=50) regarding baseline symptoms before and one month after the intervention (P>0.05). While, they were significantly different three months after the intervention (P<0.05). The mean pure tone air and bone conduction were not significantly different between the control and the intervention group before and three months after the intervention at 0.5,1,2 and 4 kHz (P>0.05). The mean pure tone air conduction was not significantly different between the two groups before and three months after the intervention at 6 kHz (P>0.05).

**Conclusion:** A three-month treatment with Neurotec Capsules beside patient education can effectively control symptoms of patients with tinnitus.

## Introduction

Tinnitus or ringing in the ear is defined as the conscious perception of sound in the ear or head in the absence of an external stimulus which is recognized in about 10-15% of people ^1,2^. About one to two percent of world population suffer from highly disabling presence of ear noise and developing comorbid conditions such as depression, demoralization and social withdrawal ^3^.

It has been shown that various social factors including low education level, poor income, and some types of occupational activities increasing the noise exposure, lead to development of tinnitus ^1^. Furthermore, medical conditions such as trauma, hearing loss, or ototoxic treatments are associated with development of tinnitus. Tinnitus is not considered a disease, but rather it is mainly considered as symptom and presentation of underling medical conditions ^4^.

The exact etiology of tinnitus is not well understood. Traditionally, tinnitus has been considered as ontological disorder. However, advancement in neuroimaging and animal models have demonstrated that it could be associated with neuronal interactions. Enhanced neuronal firing rate, increased neuronal synchrony, altered tonotopic organization of brain pathway, as well as alteration in non-auditory brain areas are considered as possible mechanisms involved in development of tinnitus ^2^.

So far, there is no available medication that could completely resolve the symptoms of tinnitus and most available treatments help patients to cope with tinnitus and increase their quality of life ^5^. In addition to pharmacotherapy, other treatments such as cognitive and behavioral therapy, sound therapy, music therapy, acupuncture, and hearing aids have been suggested for improvement of tinnitus ^4, 6^. Herbal medication such as Ginko Biloba and Gushen Pian (Chinese herbal medication) are considered as an alternative treatment for tinnitus ^6^. However, there are not enough and well-designed study to recommend these medications for treatment of tinnitus.

Neurotec® is a medication of herbal origin which contains extracts of dog-rose (*Rosa Canina*), common nettle (*Urtica Dioica*) and tansy (*Tanacetum Vulgare*). Neurotec® is an IFDA approved medication for treatment of peripheral neuropathies in diabetic patients and exerts its action through improving neural message conduction. According to a previous studies, combination of herbal extracts including *Rosa Canina, Urtica Dioica* and *Tanacetum Vulgare*, can significantly improve motor function of cerebral ischemic rats and has a protective effect against ischemic brain injury ^7^. In addition, since Neurotec® ingredients have improved spatial learning and memory in rat models of Alzheimer disease, its anti-oxidant and anti-inflammatory effects can be used as a neuroprotective herbal combination ^8^.

Considering all the above mentioned points and according to neuroprotective role of Neurotec ingredients, we hypothesized that it can improve auditory nerve function resulting in better enviromental sound perception and consequent reduced tinnitus symptoms. In this study, we used a standardized dose of Neurotec in a double-blinded randomized clinical trial to determine whether it can be effective for management of tinnitus symptoms.

## Materials and methods

Following registration at ethics committee of Baqiyatallah University of Medical Sciences (Ref. No: IR.BMSU.REC.1395.176) and Iranian Registry of Clinical Trials (Ref. No: IRCT2017042817413N23), this double-blind randomized clinical trial was conducted on patients with tinnitus, who attended otolaryngology clinic of Baqiyatallah Hospital between 2015 and 2016. All participants provided written informed consent—this study ad-hered to the principles of the Declaration of Helsinki. Figure 1 shows a flowchart of the trial.

**Figure 1.**
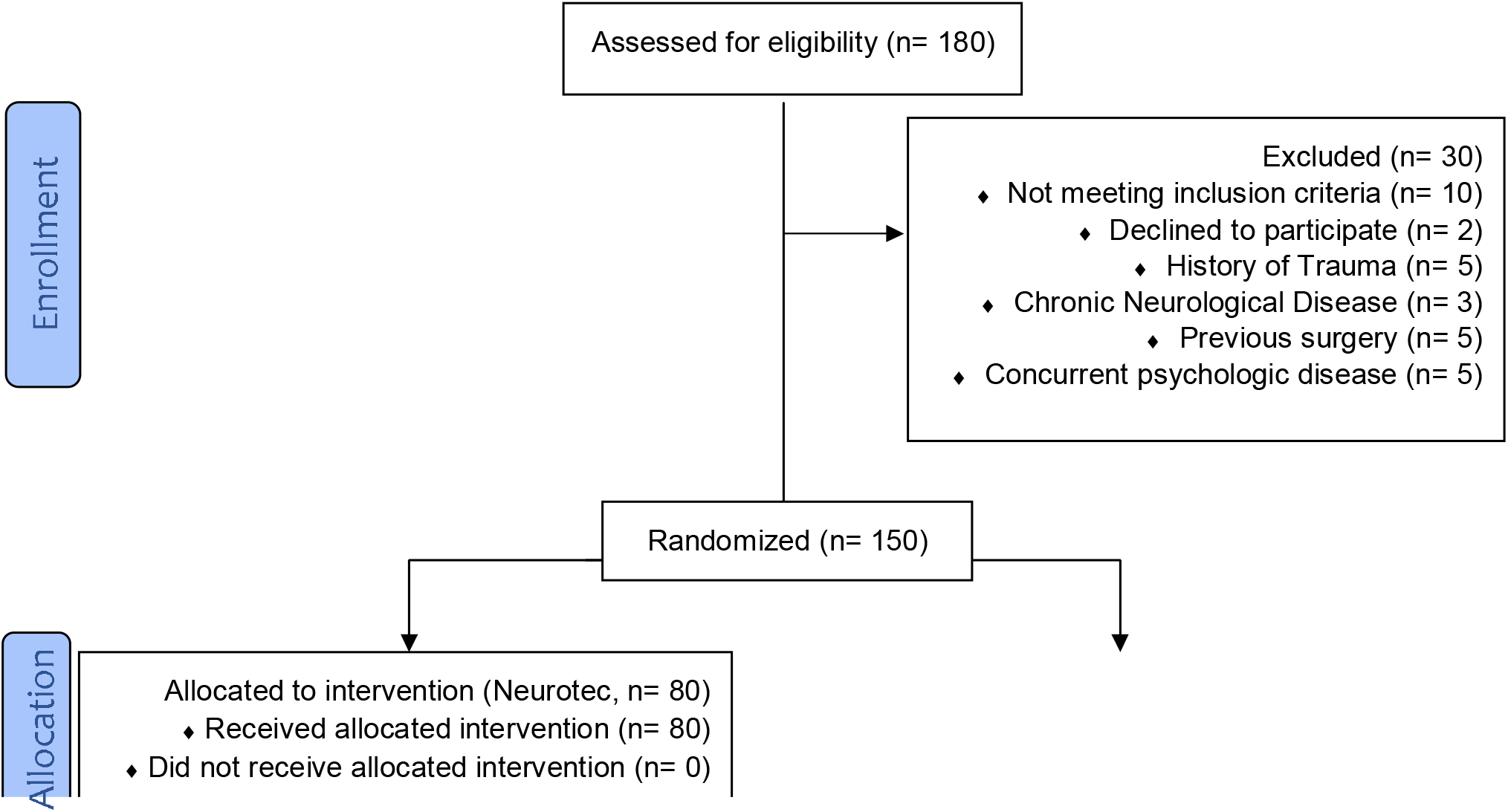
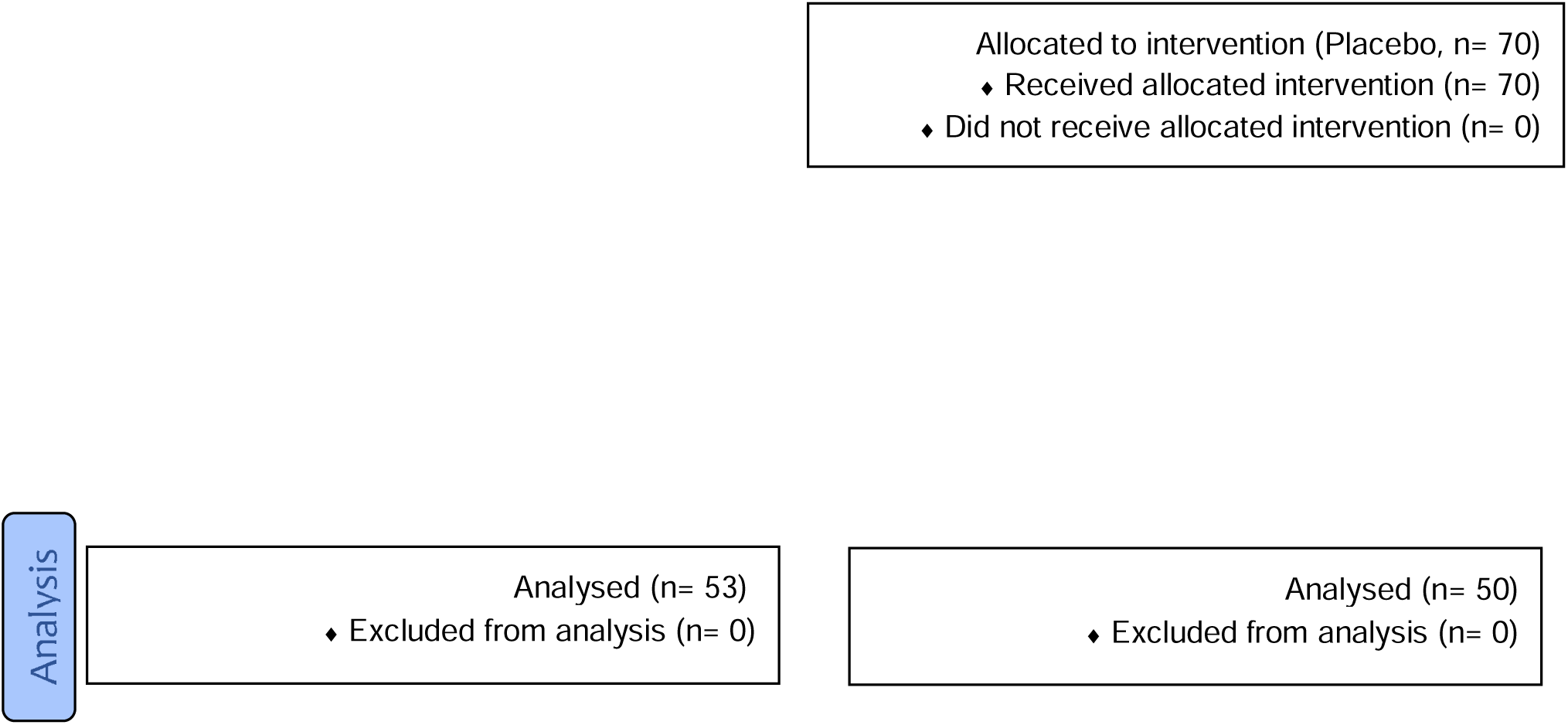
Study flowchart.

Patients were assessed for eligibility by an otolaryngologist. Patients with chief complaint of subjective tinnitus for at least 6 months and those above 19 years of age were included in the study. Patients over 75 or below 19 years of age, pregnant women, those with chronic or acute neurological diseases, concomitant psychiatric illnesses, pulsatile tinnitus and underlying conditions such as epilepsy, multiple sclerosis, and history of severe head and neck trauma or ear surgery were excluded from the trial. In addition, patients should not have taken any medication for treatment of tinnitus within at least one month prior to intervention. Patients were randomly allocated to intervention and control groups using random number table. Patients in intervention group were treated with Neurotec 100 mg capsules (BID) for three months. Patients in control group received placebo capsules, which had exactly the same shape and packaging with Neurotec 100 mg capsules, for the same duration. Patients in both groups received primary tinnitus treatments including reassurance and necessary trainings. Placebo capsules were provided by Rose Pharmed®, the Neurotec manufacturer in Iran.

Patients in both groups underwent pure tone audiometry (PTA) before and three months after intervention. Auditory threshold of patients was measured at 0.5, 1, 2, 4 and 6 KHz frequencies.

Validated Persian version of Tinnitus Handicap Inventory (THI) questionnaire was administered to patients in both groups prior to, one month and three months after intervention ^9^.

A zero-to-ten visual analog scale (VAS) was used to evaluate tinnitus loudness, daily annoyance, disturbance in daily life, daily tinnitus perception, sleep disturbance and mood alteration.Data were analyzed using Statistical Package for Social Sciences (IBM Corp. Released 2011. IBM SPSS Statistics for Windows, Version 20.0. Armonk, NY: IBM Corp).Descriptive analysis was performed using mean and standard deviation as well as percentages and frequencies. Kolmogorov-smirnov test was used to check the normal distribution of data. Chi-square test was used to compare the categorical variables and Fisher’s exact test if they were not normally distributed. Comparison between groups was performed using t-test, ANOVA, Kruskal-Wallis, Wilcoxon-signed rank and Mann-Whitney U test. A p-value of less than 0.05 was considered as statistically significant.

## Results

Finally, a total of 103 patients (53 cases in intervention and 50 in control group) with a mean age of 51.33±13.91 years underwent analysis. Among the patients, 69 (67%) were male with a mean age of 49.39±15.02 and 34(33%) were female with a mean age of 55.26±10.47 (p=0.024).

There was no statistically significant differences between the mean age of patients in intervention (52.96±14.64) and control (49.60±13.02) groups (p =0.222).

The intervention group included 35 (66%) and the control group included 34 (68%) male patients. The gender distribution was not significantly different between the two groups (p=0.834). Twenty five (24.3%) patients had right side, 40 (38.8%) had left side and 38 (36.9%) patients had bilateral tinnitus.

Among the patients, 36 patients (35%) had tinnitus symptoms for less than a year; 28 patients (27.2%) had symptoms for one to three years; 16 patients (15.5%) had symptoms for three to eleven years and 23 patients (22.3%) had symptoms for more than eleven years.

### Patients’ Symptoms

About all patients’ symptoms, such as tinnitus loudness, annoyance severity, daily life alteration, daily tinnitus perception, sleep alteration, mood disturbance and quality of life, there was no significant difference between the intervention group and the control group, before the intervention and one month after the intervention. While there was a significant difference between the intervention group and the control group, three months after the intervention, as shown in **Table 1**.

**Table 1:**
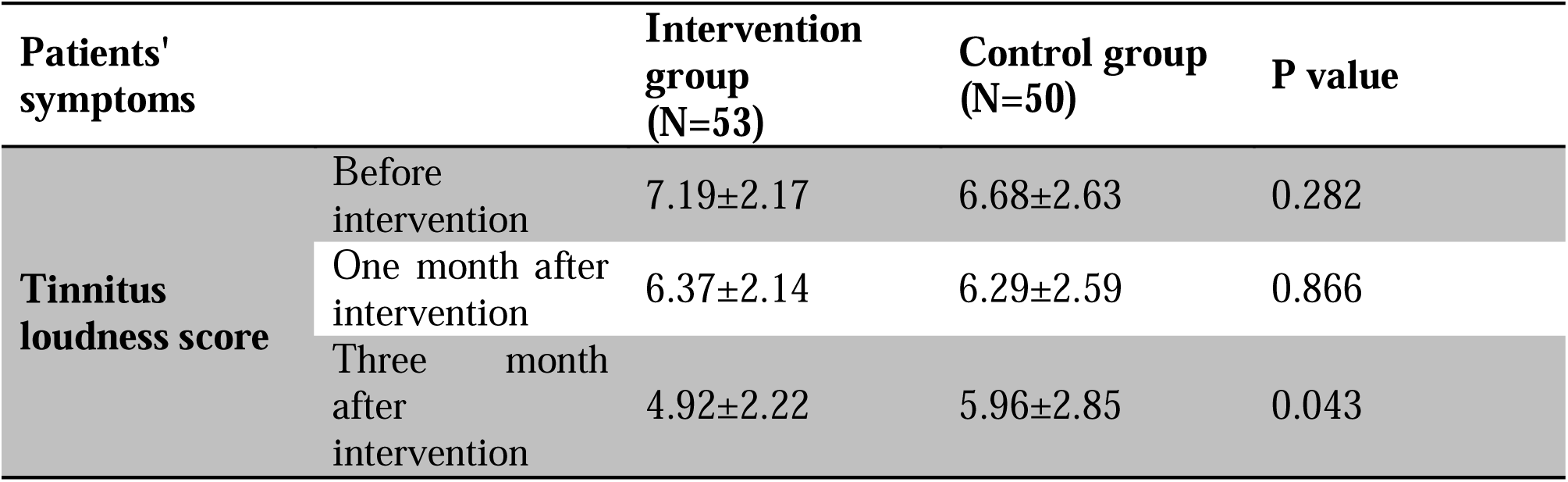
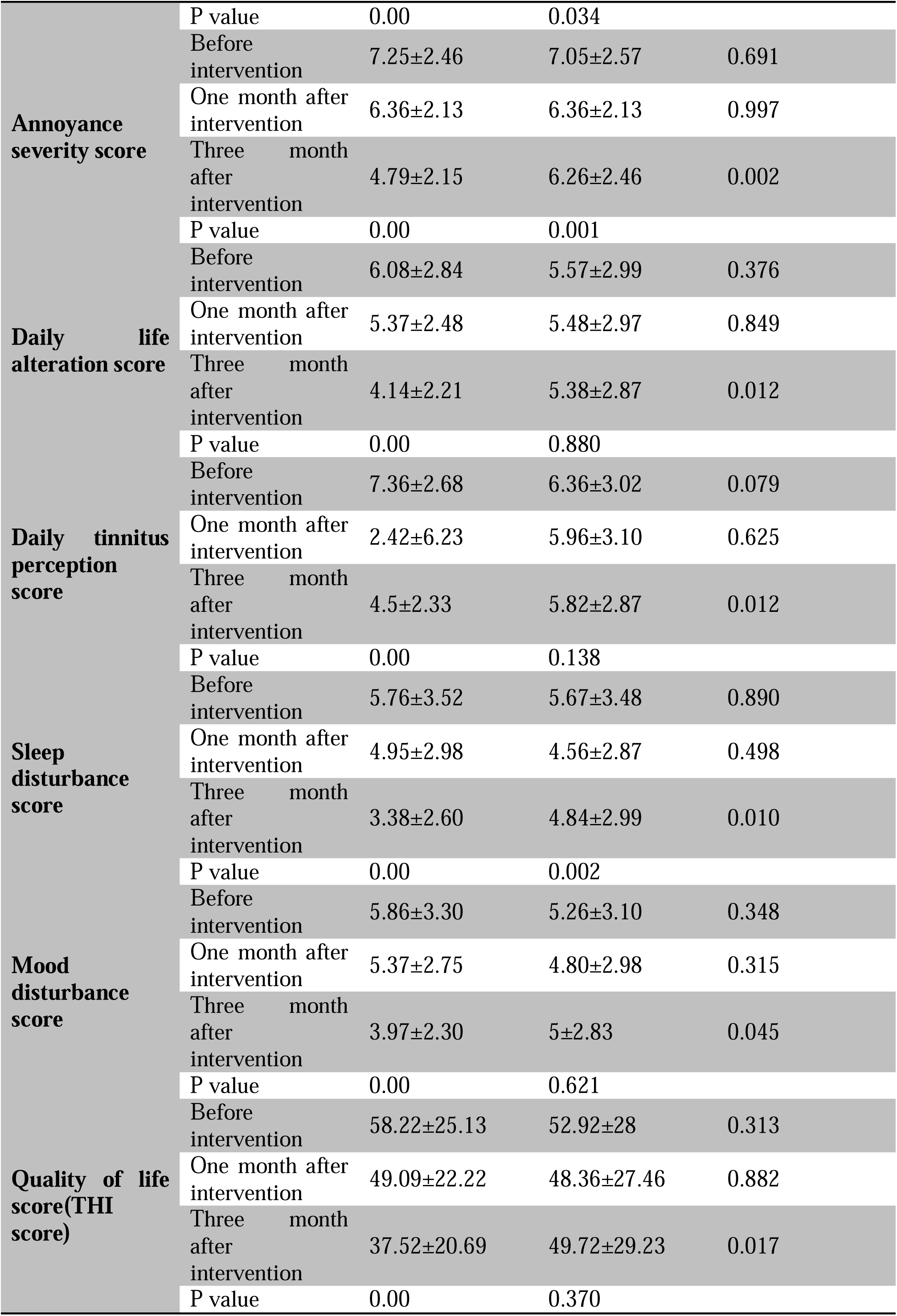
Mean THI and VAS scores before, one moth and three months after intervention

### Audiometric Test

Pure tone audiometry thresholds were analyzed in two frequency groups: low frequency (LF: 0.5, and 1 kHz) and high frequency (HF: 2, 4, and 6 kHz).

The pure tone air conduction average was not significantly different between the control group and the intervention group before the intervention and three months after the intervention at 0.5,1,2 and 4 kHz. The pure tone bone conduction average was not significantly different between the control group and the intervention group before the intervention and three months after the intervention at 0.5,1,2 and 4 kHz. The pure tone air conduction average was not significantly different between the control group and the intervention group before the intervention and three months after the intervention at 6 kHz as shown in **Table 2**.

**Table 2:**
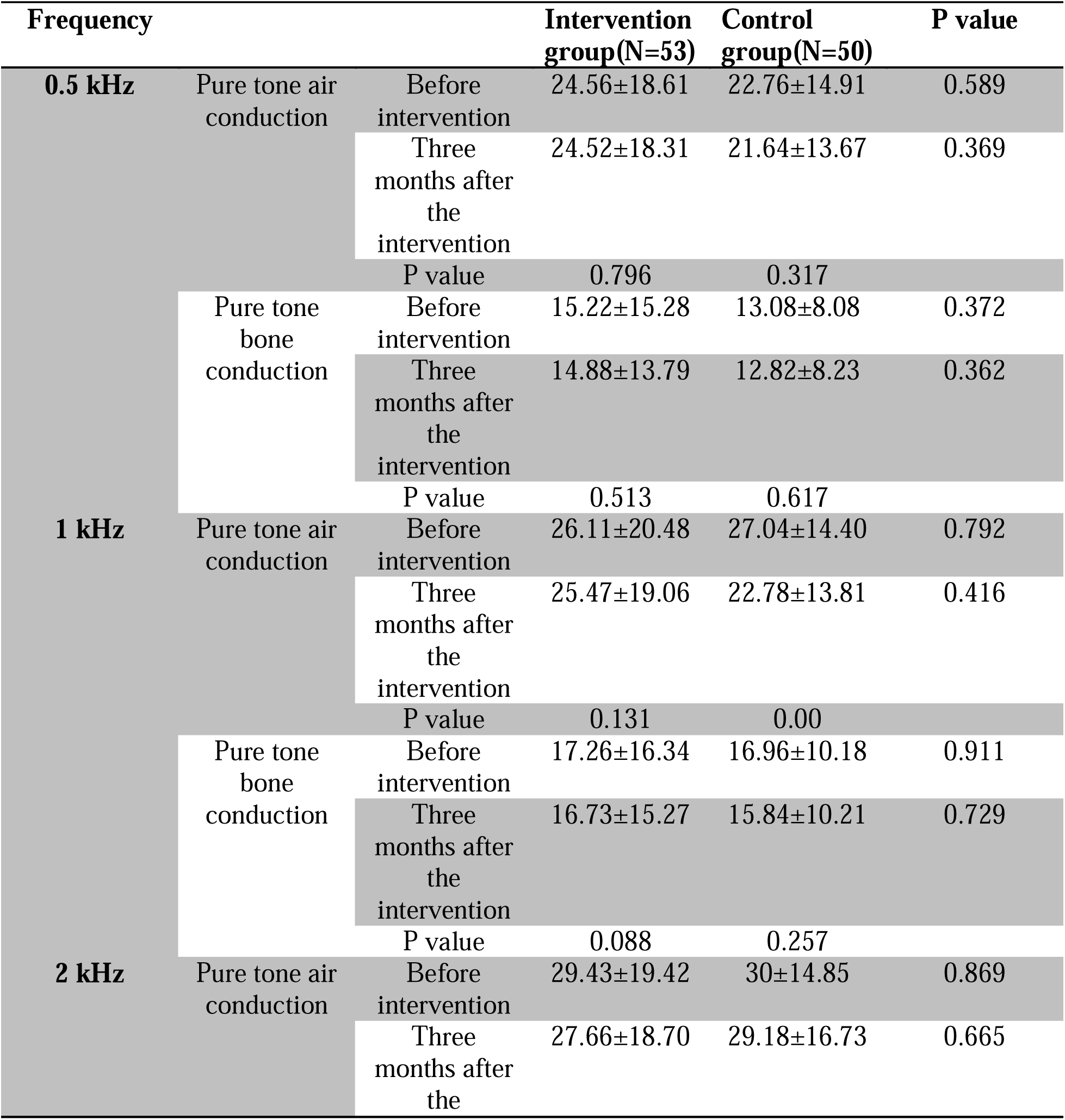
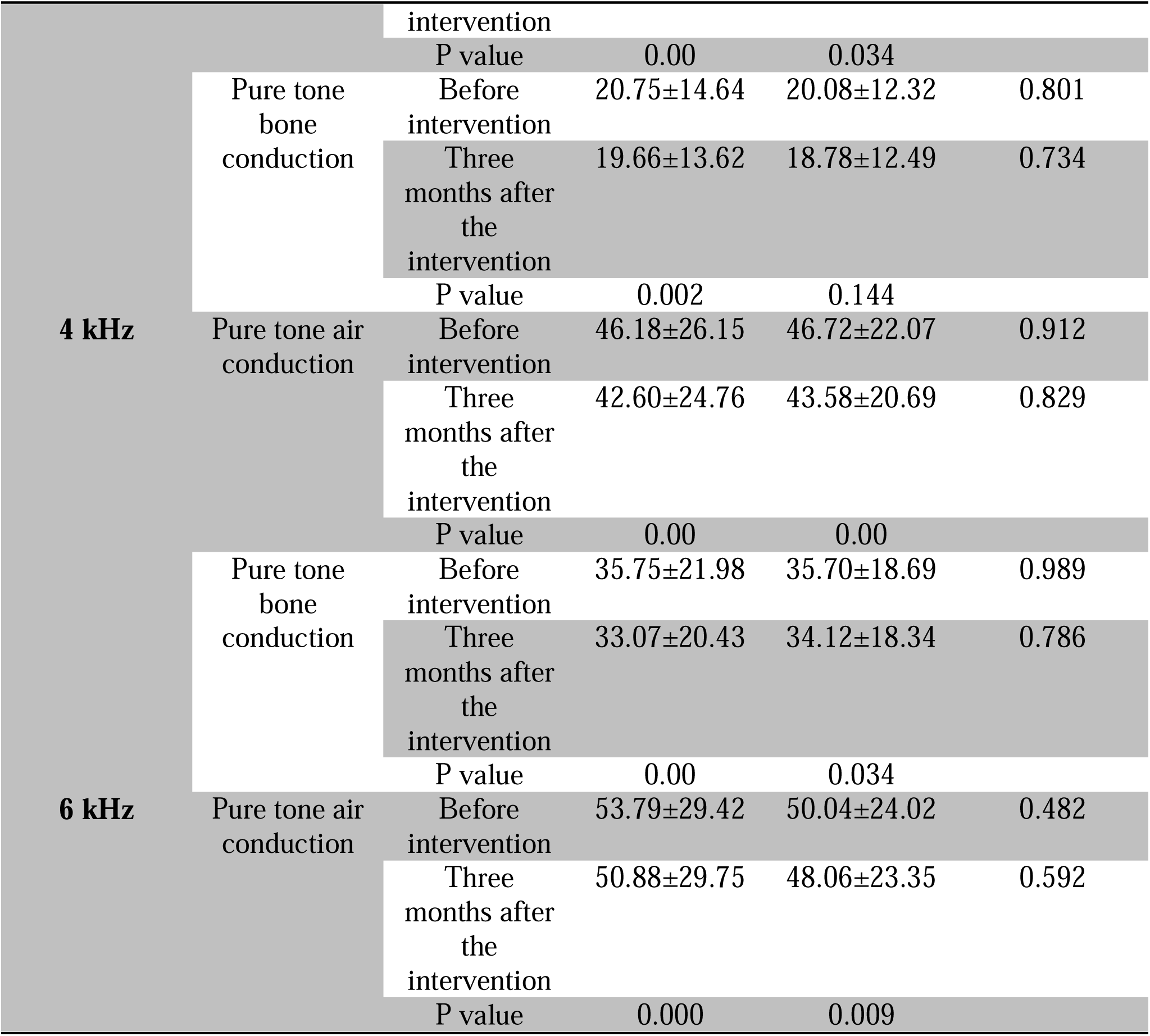
Pure tone audiometry results before and after the 3^rd^ month of intervention.

## Discussion

A variety of herbal extracts have been so far proposed for management of tinnitus; from Ginkgo biloba (Jinko) to garlic and *Yoku-kan-san* ^10^. To the best of our knowledge this is the very first study which has assessed the therapeutic effect of Neurotec (herbal combination of *Rosa Canina, Urtica Dioica* and *Tanacetum Vulgare*) on symptoms of patients with tinnitus.

We found that a three-month treatment with Neurotec beside baseline conservative treatments, significantly decreased scores of tinnitus loudness, annoyance severity, daily life alteration, sleep disturbance, daily tinnitus perception and mood disturbance in intervention group. On the other hand, control group-which was treated only by baseline educational counselling and placebo-showed the same results except for mood disturbance, daily tinnitus perception and daily life alteration scores. This is in line with results of a recently published meta-analysis in which Xiang et al., concluded that educational counseling alone is as effective as other psychological therapies for improving tinnitus and related problems ^11^. In addition, we realized that these improvements were significantly higher for the intervention group. Although patients in intervention group showed a remarkable decrease in THI score (quality of life) after three months of treatment, but those in control group experienced no significant decrease in this score.

For intervention group, pure tone air and bone conduction were significantly improved at 2, 4 and 6 kHz frequencies, after three months of treatment with Neurotec. Pure tone bone conduction did not show any significant improvement in control group at all the frequencies. However, air conduction was significantly improved in all the frequencies except 0.5 kHz.

Ghasemi et al. in an animal study concluded that Urtica Dioica extract has positive effects on learning abilities and memory impairments in wistar rats. They explained this as protective characteristic of Urtica Dioica against oxidative damage of brain tissues and acetyl choline esterase (AChE) activity ^12^. In another animal study, Patel et al. reported that Urtica Dioica can remarkably reverse diabetes chronic complications such as central and peripheral neuropathies in adult Swiss albino mice ^13^. A recently conducted animal study, has reported that Urtica Dioica extracts can effectively improve neuro-inflammation and Alzheimer-like phenotypes in rats ^14^.

Neuroprotective effects of Rosa Canina has been evaluated in various settings as in Erfani et al. study which resulted in cognitive and memory improvements in heat stress-exposed rats^15^. Assessing diabetic albino mice, Farajpour et al. showed that hydro-alcoholic extract of Rosa Canina improves memory impairment through modulation of oxidative stress ^16^. In 2016, Daneshmand et al. proved the neuroprotective effects of Neurotec ingredients on rat model of sporadic Alzheimer’s disease ^17^. Setarud (IMOD™) is an immunomodulatory drug consisted from herbal extracts of Rosa Canina, Tancetum Vulgare and Urtica Dioica. In an animal study, vafaee et al. showed that setarud has a neuroprotective effect against brain ischemia in rat models ^18^. As mentioned before, Neurotec is a safe and IFDA-approved medication for neuropathic pains, especially in diabetic patients.

Pathologies from the ear canal to the auditory cortex, may be responsible for tinnitus perception ^19^. Auditory neural plasticity is one of the considered mechanisms, which knows cochlear deafferentation as trigger, and subsequent changes in central nervous system as the cause for maintenance of tinnitus perception ^20^. We hypothesized that these neuroprotective features of Neurotec, improve cochlear nerve function and neural conduction; thus, this improvement results in better perception of environmental sounds and consequently decreases tinnitus perception. This is also the rational behind sound therapy by which, low- to moderate-level sound stimulation reverse the role of auditory depriviation in inducing tinnitus and hyperacusis ^21^.

Apart from herbal and chemical medications, a wide range of medical and non-medical treatment modalities have been evaluated for management of tinnitus; from non-invasive interventions such as yoga and meditation, sound therapy and smartphone applications to minimally- and invasive interventions like acupuncture and surgical neuromodulation ^21-25^. Despite all these efforts, no curative treatment has been so far developed for Tinnitus. It seems that patients may take benefits from an intellectual combination of these treatments^26^. On the other hand, herbal medications and extracts are an appropriate alternative therapy as they are both more tolerated by patients and have less side effects^26^.

The present study has some limitations. We had loss to follow ups in both intervention and placebo groups which may be attributable to nonsensible effects of treatment for patients, especially in initial weeks and month.

In conclusion, we found that a three-month treatment with Neurotec capsules accompanied by educational counselling is of benefit for managing symptoms in patients with chronic tinnitus. Further studies with a larger sample size and also longer duration of follow up are recommended.

## Data Availability

All data produced in the present work are contained in the manuscript

## Ethical statement

This study was approved by the Baqiyatallah University of Medical Sciences (Ref. No: IR.BMSU.REC.1395.176). All participants provided written informed consent—this study adhered to the principles of the Declaration of Helsinki.

## Data availability

The data for this article will be available upon request.

